# Infertility treatment and postpartum depressive symptoms

**DOI:** 10.1101/2025.04.28.25324402

**Authors:** SL Stone, H Diop, E Declercq, MP Fox, HJ Cabral, LA Wise

## Abstract

**BACKGROUND:** Infertility treatment (IFT) increases the likelihood of multiple gestations, which are associated with postpartum depressive symptoms (PDS). There has been little study of the association between successful IFT and risk of PDS, accounting for multiple gestations.

**METHODS:** Using representative data from the Massachusetts Pregnancy Risk Assessment Monitoring System (PRAMS) 2007-2010, we evaluated the association between self-reported IFT use and PDS. We categorized IFT as: fertility-enhancing drugs (FD), donor insemination/intrauterine insemination (DI/IUI), or assisted reproductive technology (ART) including in-vitro fertilization. We defined PDS as the report of ‘always’ or ‘often’ to any depressive symptoms. Modified Poisson regression models were used to estimate risk ratios (RRs) and 95% confidence intervals (CIs), controlling for socioeconomic status, mode of delivery, multiple gestation, neonatal risk factors and prior mental health care use.

**RESULTS:** Among 3,600 participants, 11.4% reported IFT (FD=5.9%, DI/ IUI=2.5%, ART=4.8%). Compared with non-users of IFT who delivered singletons, RRs for PDS were 1.32 (95% CI: 0.97-1.80) for IFT users who delivered singletons, 2.61 (95% CI: 1.58-4.01) for non-users of IFT who delivered multiples, and 1.06 (95% CI: 0.45-2.47) for IFT users who delivered multiples. Among mothers of singletons with Caesarean deliveries, IFT use was associated with an increased risk of PDS, RR=1.56 (95% CI: 1.03-2.38).

**CONCLUSIONS:** Non-users of IFT who delivered multiples had a greater than two-fold increased risk of PDS compared with all other groups of mothers, including IFT users who delivered singletons or multiples. IFT use was positively associated with PDS risk only among mothers with Caesarean singleton deliveries.

**Summary of Abstract:** Mothers of multiples who did not use Infertility Treatments (IFT) had more than twice the risk of PDS compared with all other groups of mothers, controlling for mode of delivery.

## INTRODUCTION

Infertility affects more than 6% of married women of reproductive age.^1^ Infertility treatment (IFT) may include fertility-enhancing drugs, donor insemination and assisted reproductive technologies that manipulate both egg and sperm in the laboratory. In unsuccessful IFT, a woman must contend with the psychological distress associated with being unable to carry a pregnancy to term,^2–5^ in addition to the financial burden often associated with IFT, and the biological effects of treatment. While a long-anticipated birth is cause for relief in some mothers who use IFT, others experience depression and anxiety about the infant after the much-sought birth.^6–8^ Only recently have studies begun to explore whether the biological effects of IFT contribute to these adverse mental health outcomes.^4, 9, 10^

Postpartum depressive symptoms (PDS) include feelings of sadness and hopelessness during the year after giving birth.^11^ The prevalence of PDS in the United States ranges from 10-15% for mothers fulfilling criteria for major depressive disorder to as high as 80% for mothers with transient ‘baby-blues’.^12^ Although there is a well-established association between maternal major depressive disorder and poor infant and child development,^11, 13, 14^ other studies suggest that even moderate levels of depressive symptoms can deleteriously affect families. Untreated PDS can impair mother-infant bonding, and delay social and cognitive development in children of affected women.^11, 15–17^ Despite there being effective treatments for PDS, between 50-80% of women with at least moderate PDS do not seek help.^18–20^ Suspected risk factors for PDS include young maternal age, obstetric complications, multiple gestations, financial difficulties and a history of major depressive episodes.^6, 7, 11, 21–27^

IFT may involve increasingly invasive technologies, from ovulation-stimulating hormones to embryo implantation. Gonadotropin-releasing hormone agonists (GnRH-a), commonly used in IFT, may induce depressive symptoms via hypogonadism. Therefore, it is reasonable to consider GnRH-a as a possible mechanism by which IFT could cause PDS.^28–31^ Emerging research suggests that IFT may be associated with an increased risk of PDS, but results have been inconsistent. In a retrospective cohort study of 745 Australian mothers, those who used assisted reproductive technology to conceive were four times as likely to seek residential early-parenting services during the first twelve months after parturition —where mothers, with their infants, obtain help and education in infant care and psycho-social support—as mothers who conceived spontaneously, but this study did not account for multiple gestations, mode of delivery or birth outcomes.^32^ ^32^ In contrast, other studies which did account for multiple gestations, mode of delivery and birth outcomes found no significant differences between conception groups on psychological status during pregnancy or postpartum.^33, 34^ Possible reasons for the lack of consistency of findings across studies are differences in how multiple gestations, mode of delivery and birth outcomes such as infant birthweight, gestational age and NICU stay were treated in analysis. Because multiple gestations and Cesarean delivery are risk factors for PDS and these outcomes are more common among IFT users, they might explain the elevated risk of PDS associated with IFT.^7, 35–38^ Our study assesses the association between IFT and risk of PDS—while accounting for multiple births and mode of delivery—in a representative sample among mothers who delivered infants during 2007-2010 in Massachusetts. We also examine the association between IFT and seeking mental health help among mothers with PDS.

## METHODS

### Participants and Procedures

Study subjects were mothers with a recent birth who participated in the Massachusetts Pregnancy Risk Assessment Monitoring System (MA-PRAMS) during 2007-2010. PRAMS is a multi-state, population-based surveillance system funded by the Centers for Disease Control and Prevention (CDC) in collaboration with state health departments. PRAMS collects data on maternal experiences that occur before and throughout pregnancy as well as in early infancy.

PRAMS methodology and protocol have been published elsewhere.^39^ MA-PRAMS includes questions on maternal characteristics, pregnancy intention, infertility treatments, birth outcomes, maternal mood and health after birth. PRAMS participants are randomly-selected 2-6 months postpartum from state live birth certificate information, representing approximately 3.0% of all female MA residents delivering a live birth during the study period. To ensure adequate representation of racial/ethnic minority groups, MA-PRAMS disproportionately samples women by race/ethnicity. The survey is administered in both English and Spanish. Mothers whose pregnancy ended in stillbirth or multiple-births resulting in greater than triplets were excluded.

MA-PRAMS has a 70% weighted response rate. The study was approved by the Institutional Review Board of the Massachusetts Department of Public Health.

### Assessment of Exposures

#### PRAMS asked

“Did you receive treatment from a doctor, nurse, or other health care worker to help you get pregnant with your new baby? (This may include infertility treatments such as fertility-enhancing drugs or assisted reproductive technology) No/Yes” “If Yes: Did you use any of the following treatments *during the month you got pregnant* with your *new* baby?”

1.) “Fertility-enhancing drugs prescribed by a doctor (fertility drugs include Clomid, Serophene, Pergonal, or other drugs that stimulate ovulation)”.
2.) “Artificial insemination or intrauterine insemination (treatments in which sperm, but NOT eggs, were collected and medically placed into a woman’s body)”.
3.) “Assisted reproductive technology (treatments in which BOTH a woman’s eggs and a man’s sperm were handled in the laboratory, such as in vitro fertilization [IVF], gamete intrafallopian transfer [GIFT], zygote intrafallopian transfer [ZIFT], intracytoplasmic sperm injection [ICSI], frozen embryo transfer, or donor embryo transfer)”.
4.) “Other medical treatment (please tell us)”.

We dichotomized IFT (yes/no) and also classified IFT as: fertility-enhancing drugs (FD), donor insemination or intrauterine insemination (DI/IUI), and assisted reproduction technology (ART). Women could contribute to more than one category.

### Assessment of Outcomes

PRAMS asked mothers questions regarding mood using a Likert-like scale and are similar to questions asked on the Patient Health Questionnaire-2 (PHQ-2) depression model,^40^ an effective screening tool for depressive symptoms (Supplemental Table 1).^41, 42^ PRAMS questions were piloted by the Centers for Disease Control and Prevention. PRAMS Phase 5 (2007-2008) asked:

> “1) Since your new baby was born, how often have you felt down, depressed or hopeless, and 2) Since your new baby was born how often have you had little interest or little pleasure in doing things? Response options were: Always, Often, Sometimes, Rarely, Never.”

### PRAMS Phase 6 (2009-2010) asked

> “Below is a list of feelings and experiences that women sometimes have after childbirth. Read each item to determine how well it describes your feelings and experiences. Then write on the line the number of the choice that best describes how often you have felt or experienced things this way since your new baby was born: A) I felt down, depressed or sad. B) I felt hopeless. C) I felt slowed down.”
>
> Response options were on a Likert scale with Never =1, Rarely =2, Sometimes=3, Often =4, Always =5, for A, B, and C each.

To define PDS, we sought component questions used on the PHQ-2 to identify depression, and guidance from the CDC.^43^ For Phase 5 participants, we defined mothers as having PDS if they reported “Always” or “Often” to either question on depressive symptoms in Phase 5. As recommended by the CDC, for Phase 6 participants we summed the scores of depression symptom responses and defined mothers as having PDS if they had summed depressive symptom scores ≥10.^43^ To have greater comparability with Phase 5 participants, we also defined mothers as having PDS if they reported “Always” or “Often” to either part A or B to Phase 6 questions. We defined mothers who reported “Sometimes/ Rarely/Never” to all questions as our reference. This method yielded 63% sensitivity and 83% specificity for depression in Phase 5, and 56.8% sensitivity and 86.6% specificity in Phase 6, compared to the PHQ-2.^43^

PRAMS also asked about help-seeking behavior specifically for depression:

> “Since your new baby was born, have you asked for help for depression from a doctor, nurse or other health care provider? Yes/No.”

### Assessment of Covariates

PRAMS collected information on a wide range of variables associated with both IFT and PDS in the literature (i.e., potential confounders), including maternal age, pre-pregnancy body mass index (BMI), plurality, mode of delivery, gestational age, congenital malformations, birth weight, stay in neonatal intensive care unit (NICU), race/ethnicity, government-paid health care (during pregnancy, delivery and postnatally), marital status, education, parity, yearly family income, cigarette use in pregnancy, infant sex, length of hospital stay and mother’s pre-pregnancy mental health visits (available for PRAMS Phase 6, 2009-2010).

### Exclusions

Of the 5,899 mothers who completed PRAMS in 2007, 2008, 2009 or 2010, we excluded mothers with missing or implausible data on PDS variables (n=256), mothers whose infant had died shortly after birth before completion of PRAMS (n=40), and mothers who indicated that they wanted to be pregnant later (n=1,649) or did not want to be pregnant then or at any time in the future (n=448), because mothers with ill-timed or unintentional pregnancies may have different risks for PDS than women who planned a pregnancy. Excluded mothers were more likely to have lower education, lower family income, higher parity, and be Black non-Hispanic than mothers who remained in our analysis (data not shown). After exclusions, 3,600 mothers remained.

### Data Analysis

We used SUDAAN to account for the complex survey design of PRAMS and to obtain frequencies weighted by race/ethnicity. We used modified Poisson regression models with a robust error variance to directly estimate risk ratios (RR) and 95% confidence intervals (CI) for the association between IFT and PDS.^44^ Potential confounders that changed the effect estimate by more than 10% were retained in the final models.^45, 46^ Possible causal intermediates, identified through directed acyclic graphs, were excluded.^47, 48^

Based on these criteria, we controlled for maternal age (categories based on frequency distributions: <25, 25-29, 30-34, ≥ 35 years), education (< high school diploma, high school diploma, some college, and completed college), family income (<$15k, $15k-24.9k, $25k-49.9k, and ≥$50k), multiple gestations (singleton/multiple), mode of delivery (vaginal/Cesarean) and prior mental health visit (in Phase 6—mother reported pre-pregnancy visit for anxiety or depression (yes/no). We also considered calendar year of PRAMS, as IFT prevalence increased yearly from 2007-2010. Among mothers with PDS, we examined the association between IFT (any use; category) and seeking help, defined as “since your new baby was born, have you asked for help for depression from a doctor, nurse or other healthcare worker? Yes/No” with “no reported IFT” as our reference. Regression analyses utilized SAS version 9.3 (SAS Institute, Inc. NC).

## RESULTS

Selected baseline characteristics of mothers and their infants are presented in Table 1. Mothers reporting any IFT use were more likely to be older, married, White non-Hispanic, have higher education and family income, have diabetes, or be obese (BMI ≥ 30) and less likely to smoke than spontaneously conceiving (SC) mothers. The infants of users of IFT were more likely to be born with gestational age <37 weeks (13.5% versus 5.4%), have low birth weight (14.1% versus 5.5%) and require a NICU stay (14.7% versus 10.9%) when compared with infants of non-users of IFT. Multiple gestations occurred in 14.2% of users of IFT and 1.1% of non-users of IFT. Among multiple gestations, infants of users of IFT were less likely to be preterm (55.0% versus 61.3%), have low birth weight or very low birth weight (50.5% versus 79.6% and 0.8% versus 6.9% respectively), and require a NICU stay (38.5% versus 56.1%), compared with infants of non-IFT users.

**Table 1.**
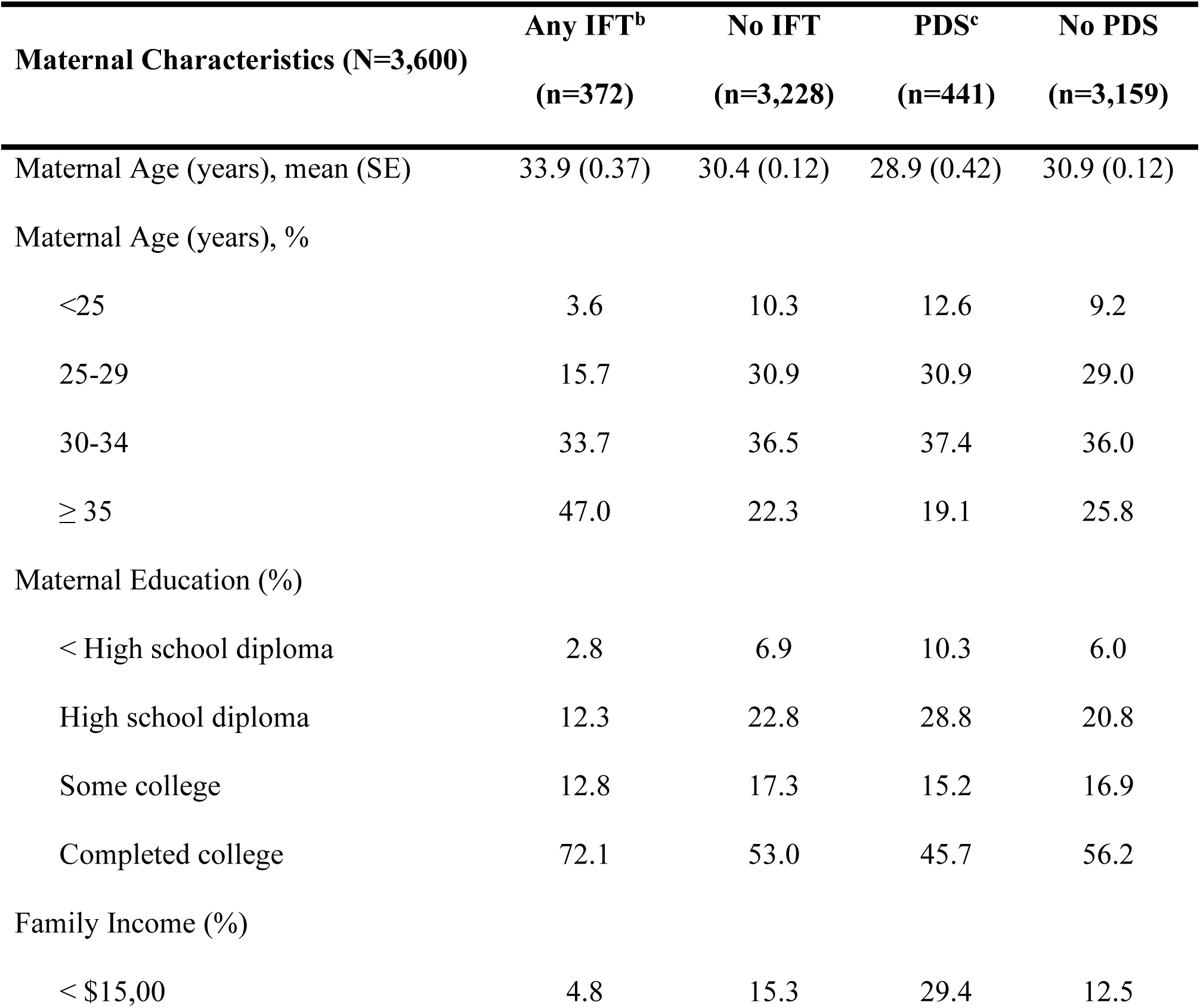
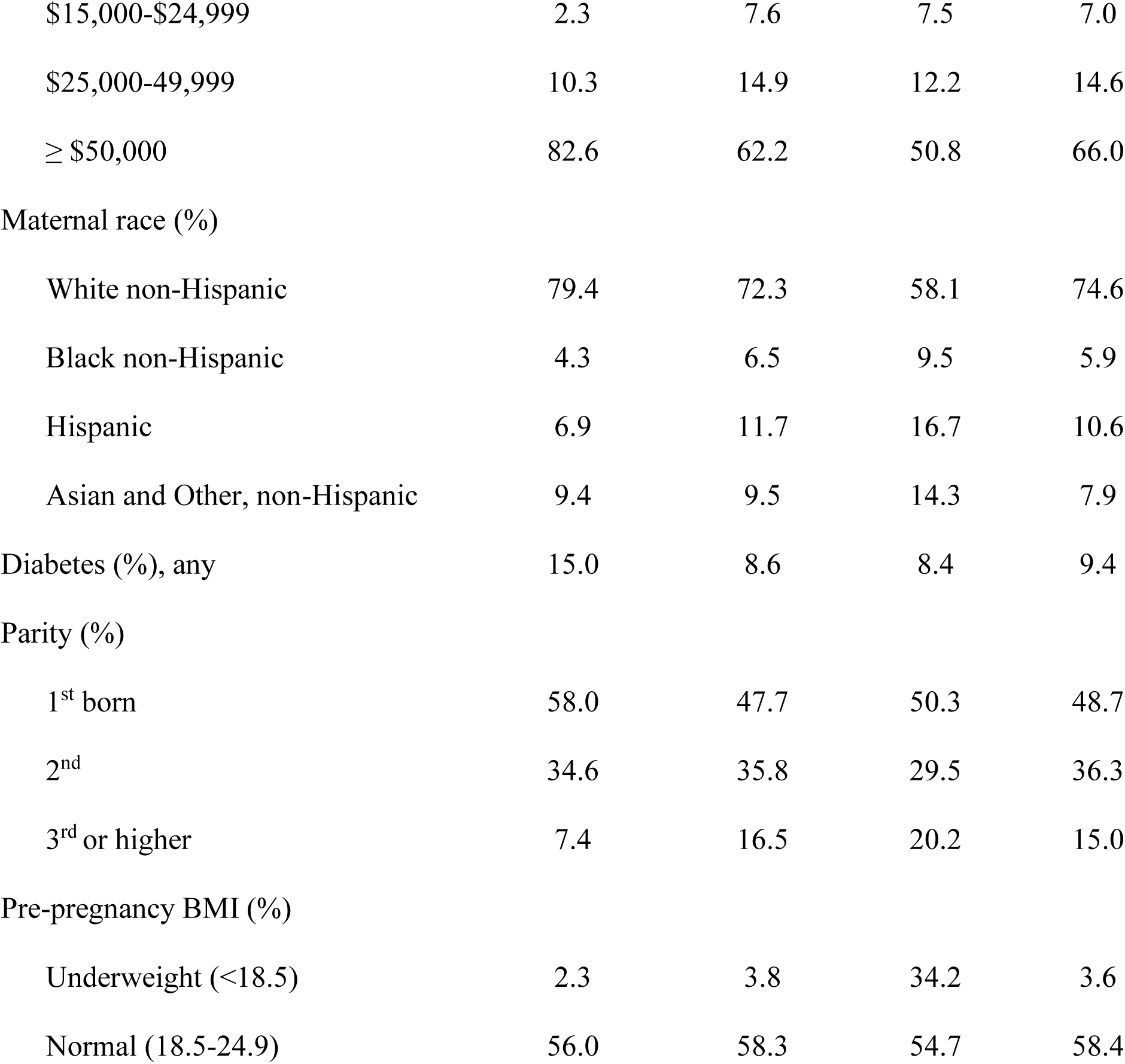
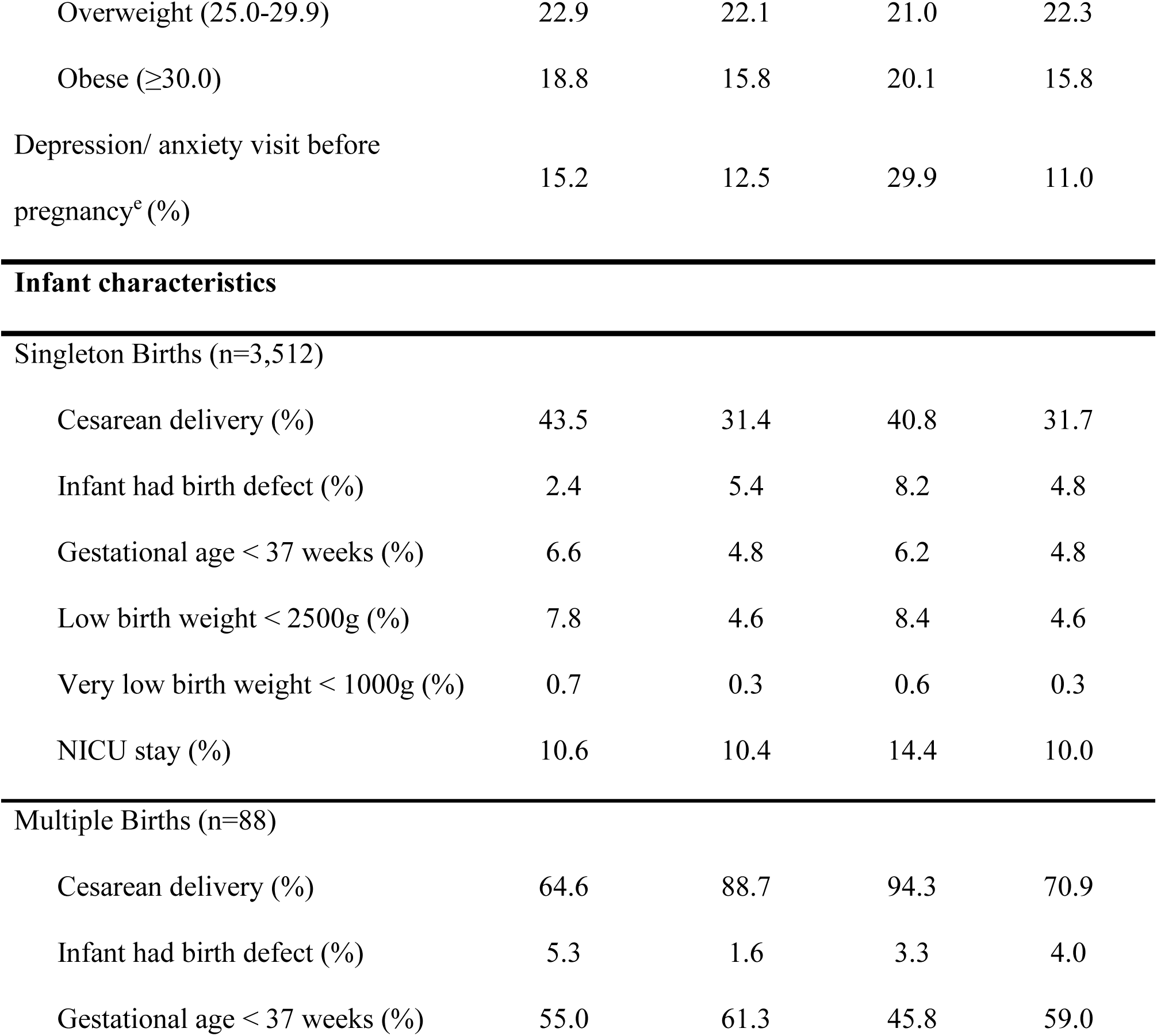
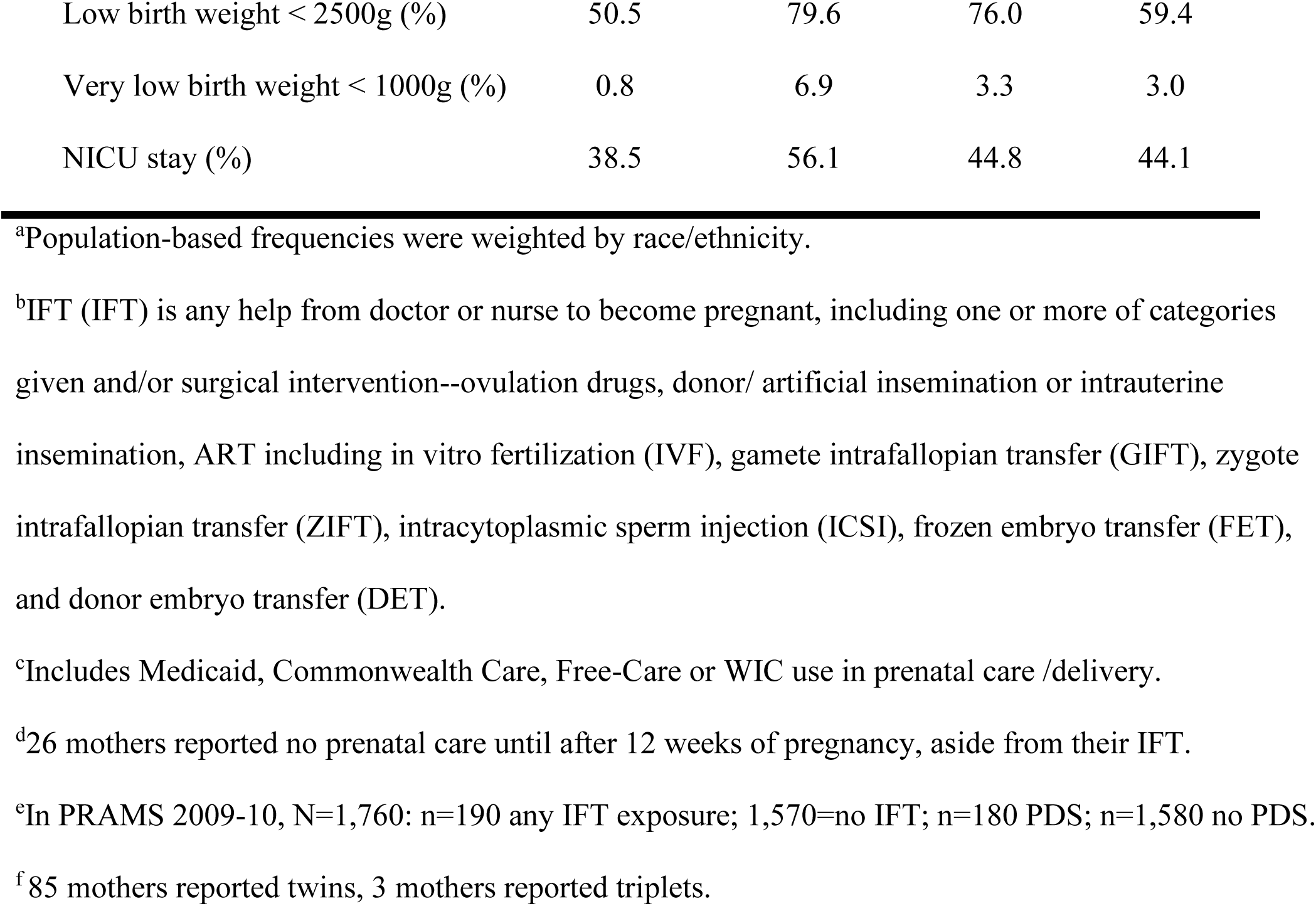
Characteristics of mothers and their infants in PRAMS-MA, 2007-2010^a^.

| **Maternal Characteristics (N=3,600)** | **Any IFT<sup>b</sup>**<br>**(n=372)** | **No IFT**<br>**(n=3,228)** | **PDS<sup>c</sup>**<br>**(n=441)** | **No PDS**<br>**(n=3,159)** |
| --- | --- | --- | --- | --- |
| Maternal Age (years), mean (SE) | 33.9 (0.37) | 30.4 (0.12) | 28.9 (0.42) | 30.9 (0.12) |
| Maternal Age (years), % |  |  |  |  |
| <25 | 3.6 | 10.3 | 12.6 | 9.2 |
| 25-29 | 15.7 | 30.9 | 30.9 | 29.0 |
| 30-34 | 33.7 | 36.5 | 37.4 | 36.0 |
| ≥ 35 | 47.0 | 22.3 | 19.1 | 25.8 |
| Maternal Education (%) |  |  |  |  |
| < High school diploma | 2.8 | 6.9 | 10.3 | 6.0 |
| High school diploma | 12.3 | 22.8 | 28.8 | 20.8 |
| Some college | 12.8 | 17.3 | 15.2 | 16.9 |
| Completed college | 72.1 | 53.0 | 45.7 | 56.2 |
| Family Income (%) |  |  |  |  |
| < \$15,00 | 4.8 | 15.3 | 29.4 | 12.5 |
| \$15,000-\$24,999 | 2.3 | 7.6 | 7.5 | 7.0 |
| \$25,000-49,999 | 10.3 | 14.9 | 12.2 | 14.6 |
| ≥ \$50,000 | 82.6 | 62.2 | 50.8 | 66.0 |
| Maternal race (%) |  |  |  |  |
| White non-Hispanic | 79.4 | 72.3 | 58.1 | 74.6 |
| Black non-Hispanic | 4.3 | 6.5 | 9.5 | 5.9 |
| Hispanic | 6.9 | 11.7 | 16.7 | 10.6 |
| Asian and Other, non-Hispanic | 9.4 | 9.5 | 14.3 | 7.9 |
| Diabetes (%), any | 15.0 | 8.6 | 8.4 | 9.4 |
| Parity (%) |  |  |  |  |
| 1<sup>st</sup> born | 58.0 | 47.7 | 50.3 | 48.7 |
| 2<sup>nd</sup> | 34.6 | 35.8 | 29.5 | 36.3 |
| 3<sup>rd</sup> or higher | 7.4 | 16.5 | 20.2 | 15.0 |
| Pre-pregnancy BMI (%) |  |  |  |  |
| Underweight (<18.5) | 2.3 | 3.8 | 34.2 | 3.6 |
| Normal (18.5-24.9) | 56.0 | 58.3 | 54.7 | 58.4 |
| Overweight (25.0-29.9) | 22.9 | 22.1 | 21.0 | 22.3 |
| Obese ($\geq 30.0$) | 18.8 | 15.8 | 20.1 | 15.8 |
| Depression/ anxiety visit before pregnancy<sup>e</sup> (%) | 15.2 | 12.5 | 29.9 | 11.0 |
| Singleton Births (n=3,512) |  |  |  |  |
| Cesarean delivery (%) | 43.5 | 31.4 | 40.8 | 31.7 |
| Infant had birth defect (%) | 2.4 | 5.4 | 8.2 | 4.8 |
| Gestational age < 37 weeks (%) | 6.6 | 4.8 | 6.2 | 4.8 |
| Low birth weight < 2500g (%) | 7.8 | 4.6 | 8.4 | 4.6 |
| Very low birth weight < 1000g (%) | 0.7 | 0.3 | 0.6 | 0.3 |
| NICU stay (%) | 10.6 | 10.4 | 14.4 | 10.0 |
| Multiple Births (n=88) |  |  |  |  |
| Cesarean delivery (%) | 64.6 | 88.7 | 94.3 | 70.9 |
| Infant had birth defect (%) | 5.3 | 1.6 | 3.3 | 4.0 |
| Gestational age < 37 weeks (%) | 55.0 | 61.3 | 45.8 | 59.0 |
| Low birth weight < 2500g (%) | 50.5 | 79.6 | 76.0 | 59.4 |
| Very low birth weight < 1000g (%) | 0.8 | 6.9 | 3.3 | 3.0 |
| NICU stay (%) | 38.5 | 56.1 | 44.8 | 44.1 |

<sup>a</sup>Population-based frequencies were weighted by race/ethnicity.
<sup>b</sup>IFT (IFT) is any help from doctor or nurse to become pregnant, including one or more of categories given and/or surgical intervention--ovulation drugs, donor/ artificial insemination or intrauterine insemination, ART including in vitro fertilization (IVF), gamete intrafallopian transfer (GIFT), zygote intrafallopian transfer (ZIFT), intracytoplasmic sperm injection (ICSI), frozen embryo transfer (FET), and donor embryo transfer (DET).
<sup>c</sup>Includes Medicaid, Commonwealth Care, Free-Care or WIC use in prenatal care /delivery.
<sup>d</sup>26 mothers reported no prenatal care until after 12 weeks of pregnancy, aside from their IFT.
<sup>e</sup>In PRAMS 2009-10, N=1,760: n=190 any IFT exposure; 1,570=no IFT; n=180 PDS; n=1,580 no PDS.
<sup>f</sup>85 mothers reported twins, 3 mothers reported triplets.

Frequencies of IFT were 2.5% for DI/IUI, 4.8% for ART, and 5.9% for FD, with a combined prevalence of IFT use of 11.4%. Overall, 441 mothers (9.5%) reported PDS. Less than 40.0% of mothers with PDS reported seeking help (Supplemental Table 2). 9.1% of non-users of IFT with singletons reported PDS compared to 12.3% of users of IFT with singletons (Table 2). Among multiple gestations, 6.4% of users of IFT reported PDS, compared with 22.6% among non-users of IFT mothers of multiples. Multivariate RRs for PDS were 1.26 (95% CI: 0.94-1.69) for users of IFT with singletons, 1.08 (95% CI: 0.50-2.32) for users of IFT with multiples, and 2.38 (95% CI: 1.42-3.98) for non-users of IFT with multiples, all compared with non-users of IFT with singletons. Applying the relative excess risk for interaction formula to our data, we observed that 19.4% fewer mothers with multiple gestations and IFT had PDS than would have been expected based on the additive effects of IFT and multiple gestations alone, suggesting substantial interaction between the two. After adjusting for maternal age and the use of other infertility treatments, among mothers of singletons, FD, DI/IUI and ART use each showed positive associations with PDS risk compared with non-users of IFT, although the 95% CIs included the null. Mothers of multiples who used FD, DI/IUI and ART showed no evidence of an association with PDS (Supplemental Table 3).

**Table 2.**
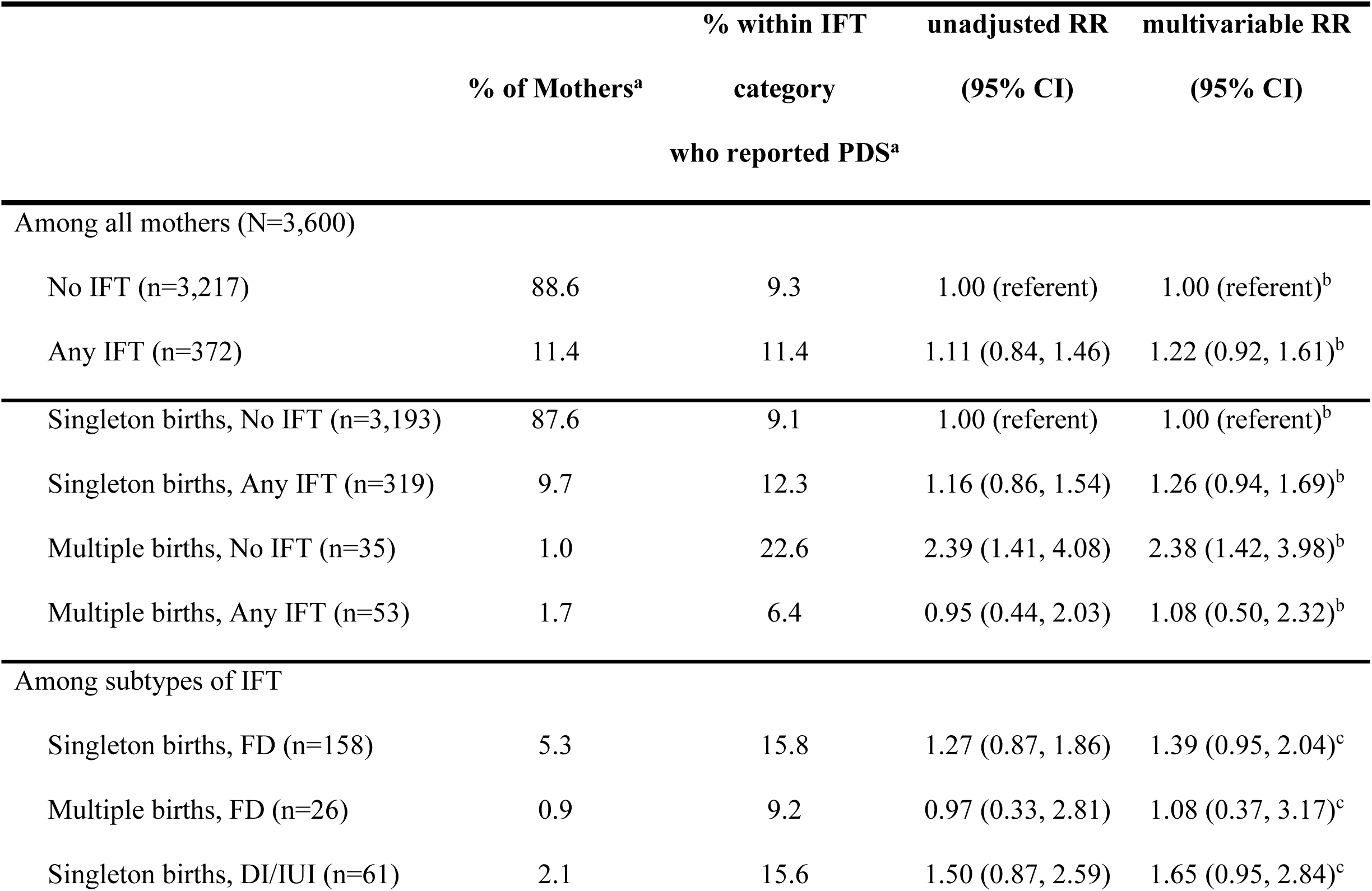
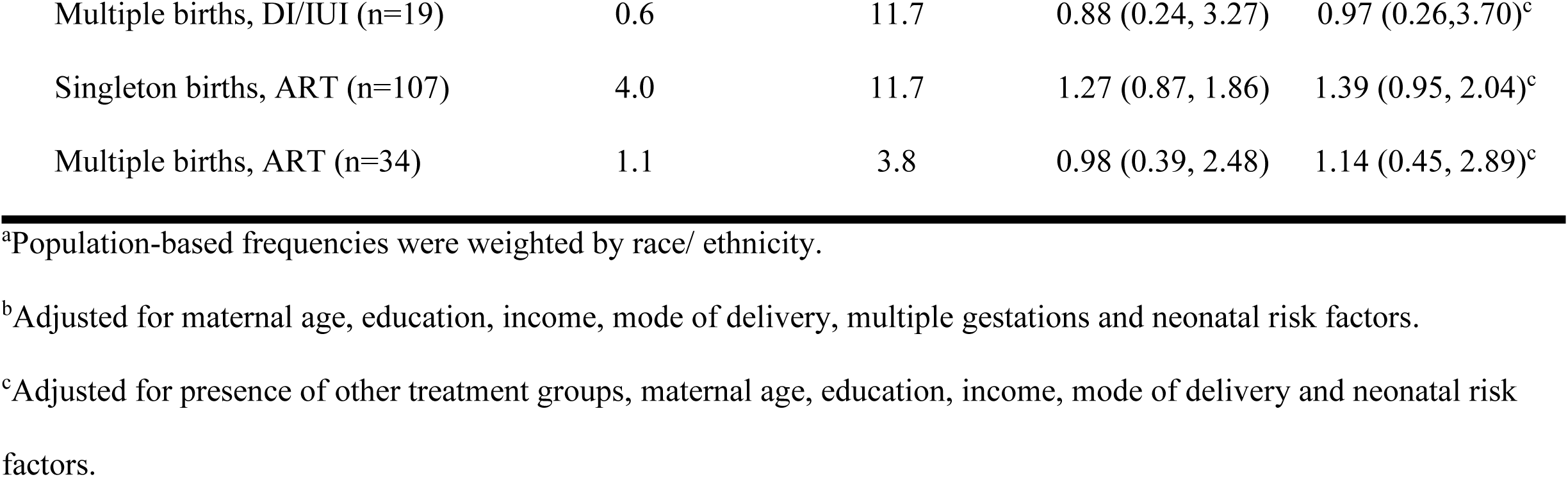
IFT and the risk of PDS among 2007-2010 PRAMS-MA mothers, by IFT category and plurality.

Stratification by mode of delivery showed differing effects (Table 3). Among 2,373 mothers who had a vaginal delivery, users of IFT with multiples showed a decrease in the risk of PDS, compared with both non-users of IFT with singletons and non-users of IFT with of multiples, although the CIs were wide. Among 1,210 mothers who had a Cesarean delivery, non-users of IFT with multiples showed an increase in risk of PDS, compared to non-users of IFT with singletons, RR=2.19 (95% CI: 1.27, 3.78). Users of IFT with singletons who had a Cesarean delivery showed an increased risk of PDS when compared to non-users of IFT with singletons, with RR=1.50 (95% CI: 0.99, 2.25). Users of IFT with multiples who had a Cesarean delivery also showed an increased risk of PDS, but these values were imprecise.

**Table 3.** IFT and the risk of PDS among 2007-2010 PRAMS-MA mothers, by plurality and delivery^a^.

|  |  | **% within IFT** | **unadjusted RR** | **multivariable** |
| --- | --- | --- | --- | --- |
|  | **% of Mothers<sup>b</sup>** | **category who** | **(95% CI)** | **RR<sup>b</sup>** |
|  |  | **reported PDS<sup>b</sup>** |  | **(95% CI)** |
| **Among mothers with vaginal births** |  |  |  |  |
| (N=2,373) |  |  |  |  |
| Singleton births, No IFT (n=2,162) | 90.6 | 8.3 | 1.00 (referent) | 1.00 (referent) |
| Singleton births, Any IFT (n=188) | 8.3 | 8.9 | 0.99 (0.65, 1.51) | 1.07 (0.77, 1.62) |
| Multiple births, No IFT (n=6) | 0.2 | 9.7 | 1.48 (0.25, 8.91) | 1.53 (0.30, 7.90) |
| Multiple births, Any IFT (n=17) | 0.9 | 1.4 | 0.52 (0.08, 3.52) | 0.55 (0.08, 3.70) |
| **Among mothers with Cesarean births** |  |  |  |  |
| (N=1,210) |  |  |  |  |
| Singleton births, No IFT (n=1,015) | 81.6 | 11.1 | 1.00 (referent) | 1.00 (referent) |
| Singleton births, Any IFT (n=131) | 12.6 | 16.8 | 1.30 (0.87, 1.95) | 1.50 (0.99, 2.25) |
| Multiple births, No IFT (n=29) | 2.7 | 24.3 | 2.30 (1.31, 4.04) | 2.19 (1.27, 3.78) |
| Multiple births, Any IFT (n=35) | 3.1 | 9.4 | 1.06 (0.46, 2.42) | 1.32 (0.57, 3.06) |

<sup>a</sup>Delivery mode data missing for 17 mothers.
<sup>b</sup>Population-based frequencies were weighted by race/ethnicity.
<sup>c</sup>Adjusted for maternal age, education, income, mode of delivery, multiple gestations and neonatal risk factors.

Mothers with a history of mental health conditions are at a greater risk of PDS.^12, 22–25, 49^ If a mother’s mood is related to changes in gonadal levels that also affect her fertility, then a history of mood disorder meets our criteria for a potential confounding variable. However, in our PRAMS dataset, data collection on prior mental health visits first began in 2009. 1,760 mothers were included in our analysis of PRAMS phase 6 2009-2010. When we adjusted for mother’s prior mental health visits, the RR among non-users of IFT with multiples was 3.38 (95% CI: 1.64-7.00) and for users of IFT with singletons RR =1.52 (95% CI: 0.99-2.35), both compared to non-users of IFT with singletons (data not shown). Among mothers who reported no prior mental health visit, users of IFT with singletons had increased risk of PDS (RR=1.78, 95% CI: 1.09-2.90), users of IFT with multiples had RR=2.37 (95% CI: 0.94-5.99) and non-users of IFT with multiples had a substantially increased risk of PDS (RR=4.89, 95% CI: 2.60-9.21), compared with non-users of IFT with singletons. Among mothers who reported a prior mental health visit, no mothers of multiples—whether IFT-users or non-users—also reported PDS, although these represented very few women. When we examined the association between categories of IFT and PDS, our results were similar; mothers who reported no prior mental health visit who used any IFT, FD, or DI/IUI all showing a significant positive association with PDS prevalence (Supplemental Table 3).

Of the 441 mothers who reported PDS, 38.6% reported seeking help for PDS. Of the mothers who reported seeking help for ‘depressive symptoms’ only 39.5% also reported PDS. We found no evidence of an association between IFT and seeking help for PDS (Table 4).

**Table 4.** IFT and Seeking Help among 441 PRAMS mothers reporting PDS, 2007-2010.

|  | **%, of Mothers**<br>**with PDS<sup>a</sup>** | **Of those with PDS,**<br>**% Sought Help<sup>a,b</sup>** | **unadjusted RR**<br>**(95% CI)** | **multivariable RR<sup>c</sup>**<br>**(95% CI)** |
| --- | --- | --- | --- | --- |
| Singleton births, No IFT | 83.9 | 39.2 | 1.00 (referent) | 1.00 (referent) |
| Singleton births, Any IFT | 12.6 | 37.1 | 1.02 (0.63, 1.66) | 1.04 (0.64, 1.69) |
| Multiple births, No IFT | 2.4 | 40.4 | 1.39 (0.64, 3.01) | 1.34 (0.62, 2.91) |
| Multiple births, Any IFT | 1.1 | 7.6 | 0.58 (0.10, 3.48) | 0.61 (0.10, 3.92) |

<sup>a</sup>Population-based frequencies were weighted by race/ethnicity.
<sup>b</sup>128 Mothers with PDS reported seeking help, representing 38.6% of PDS mothers.
<sup>c</sup>Adjusted for maternal age, education, income, mode of delivery, multiple gestations and neonatal risk factors.

## DISCUSSION

In this population-based representative study of Massachusetts mothers who gave birth during 2007-2010, we found a modest positive association between IFT and risk of PDS overall, but confidence intervals included the null. Although multiple gestations were more frequent with IFT use and were considered an independent risk factor for PDS, we found a strong inverse association between IFT and PDS among mothers of multiples, indicating that non-users of IFT who delivered multiples had greater risk of PDS than mothers of multiples who used any IFT and for all categories of IFT. This finding was persistent across mode of delivery. Among mothers of singletons, we found a positive trend of association between any IFT and PDS, and between FD, DI/IUI and ART and PDS. Among mothers with PDS, we found little evidence of an association between IFT and seeking help for depression, regardless of plurality. Although it is possible that mothers who underwent IFT had more difficulty recognizing the symptoms of depression because of unrealistic and idolized expectations of parenting once the much-sought-after baby is born,^4, 50–52^ caution must be used in interpreting this observation due to small numbers.

Our study has strengths and limitations to note. We were able to control for a wide range of potential confounders, including the differences in association by multiple gestations. Effectively since 2007, Massachusetts has required basic health insurance for all Commonwealth residents. While mental health care coverage is mandated for all mothers, only private insurance companies are required to cover IFT.^53^ Mothers who do not have or cannot afford private health insurance are eligible for health insurance through MassHealth / Commonwealth Care, which cover mental health care, but not IFT expenses.

By using a population-sampled survey, weighted by race and ethnicity, our study was designed to be representative of all Massachusetts mothers with a recent hospital birth. PRAMS has a 70% weighted response rate. In both MA-PRAMS 2007/2008 and 2009 Surveillance Reports,^54, 55^ questionnaire respondents were comparable with the state birth population in maternal characteristics of race/ethnicity, age, language and marital status. However, in 2009, 73.1% of PRAMS respondents were US-born, and 49.0% had no previous live births compared with 69.9% and 54.2% respectively, of the MA birth population. As we had no exposure or outcome data for non-respondents, we could not directly evaluate the extent to which selection bias influenced our results. Nonetheless, if participation was lower among users of IFT and mothers who developed PDS, then our observed PR might be underestimated.

Because the majority of PRAMS mothers responded within four months post-partum, some mothers who indicated that they did not experience PDS at the time of PRAMS may have subsequently developed or recognized these symptoms.^56^ However, studies indicate that the vast majority of mothers who develop PDS develop symptoms by three months postpartum.^56–58^ This would have again led to under-ascertainment of cases and potentially a conservative RR if misclassification was influenced by exposure or if the specificity of outcome classification was less than 100%. If PDS case ascertainment was not influenced by IFT use, then misclassification would likely bias our results toward the null, suggesting a stronger RR than what we observed.

Among mothers who reported seeking help for depression (n=317), 60.5% did not meet our criteria for PDS --and yet depressive symptoms still caused sufficient impairment to these mothers such that they sought help. We did not have data on the specific reasons for IFT use, number of cycles of IFT undertaken, or of IFT use for prior births. It is possible that repeated cycles of IFT-induced hypogonadal states may have a positive association with PDS as several studies of women who were treated for gynecological problems using GnRH-a found an increase in depressive symptoms that persisted even after cessation of GnRH-a.^29, 31, 59^

We also considered the impact of misclassification of maternal prenatal mental health visits. Dependent misclassification for this covariate could bias our results, but because the RRs for models which included this covariate were similar to models without this covariate, we believe that the magnitude of bias would be quite small. Controlling for “visiting a health care worker to check for or treat depression or anxiety” did not explain the association between IFT and developing PDS.

Mothers who use IFT may have a greater sense of pregnancy-related anxiety compared with spontaneously-conceiving mothers, as addressed in a systematic review of the emerging literature on psychological aspects of early parenting after IFT by Hammarberg et al.^35^ Although this review focuses primarily on maternal anxiety about pregnancy, self-esteem, and parenting stress, an included study by Klock and Greenfeld^34^ found comparable levels of prenatal depressive symptoms between users of IFT and non-users of IFT. Possibly, users of IFT with multiples were more emotionally prepared for the burdens that accompany multiples because these mothers would have been educated to the physical and social complexities of bearing and raising twins or triplets during their IFT treatment. Klock et al present several descriptive studies which found that mothers undergoing IFT were more receptive to multiple gestations than non-users of IFT.^38^ The birth of multiples to the user of IFT may also provide a sense of relief in the completion of family size, if the mother desired to bear more than one child. The overall prevalence of IFT was 11.4% in MA-PRAMS and IFT use has been steadily increasing.^60, 61^ The prevalence of PDS in MA-PRAMS (9.5%) is consistent with national prevalence,^11, 62^ suggesting that our findings might extend to other populations with similar access to comprehensive health care.

In summary, IFT use was positively associated with PDS risk only among mothers with Caesarean singleton deliveries. Non-users of IFT who delivered multiples had a greater than two-fold increased risk of PDS compared with all other groups of mothers, including IFT users who delivered singletons or multiples. Our overall findings are reassuring to the increasing number of mothers who utilize IFT.

## Data Availability

PRAMS is a population-based survey in collaboration with state departments of health and the CDC. PRAMS data are available through an application to CDC for multi-state projects or through an application to the Massachusetts Department of Public Health for Massachusetts Data.

https://www.mass.gov/info-details/pregnancy-risk-assessment-monitoring-system-prams

