## Supplement for "Infertility treatment and postpartum depressive symptoms"

| Supplemental Table 1 Patient Health Questionnaire 2 (PHQ-2) | | | | |
| --- | --- | --- | --- | --- |
| **Over the *last 2 weeks*, how often have you been bothered by any of the following problems?** | **Not at all** | **Several days** | **More than half the days** | **Nearly every day** |
| (a) **Little interest or pleasure in doing things** | **□** | **□** | **□** | **□** |
| (b) **Feeling down, depressed, or hopeless** | **□** | **□** | **□** | **□** |

| Supplemental Table 2 | | | |
| --- | --- | --- | --- |
| **Weighted Frequencies of IFT PDS and Help-Seeking by PRAMS Phase** | | | |
|  | **All Mothers^ab^** | **2007-08**  **phase 5^ac^** | **2009-10**  **phase 6^a,d^** |
| Any IFT (IFT) (%) | 11.4 | 10.7 | 12.2 |
| FD (FD: includes Clomid, Serophene, Pergonal or other drugs that stimulate ovulation) (%) | 5.9 | 5.2 | 6.6 |
| DI/IUI (DI/IUI: sperm, but not eggs, collected and medically placed in mother) (%) | 2.5 | 2.3 | 2.8 |
| ART (ART: both sperm and eggs handled in laboratory; includes IVF, GIFT, ZIFT, ICSI, FET, DET) (%) | 4.8 | 4.1 | 5.4 |
| PDS (%) | 9.5^e^ | 9.5^f^ | 9.6^g^ |
| Sought help among those with PDS (%) | 38.6^h^ | 33.2^i^ | 44.4^j^ |
| Among those who sought help, reported PDS (%) | 39.5 | 36.3 | 42.4 |
| ^a^Population-based frequencies were weighted by race/ethnicity. | | | |
| ^b^Unweighted n=3,600, ^c^unweighted n=1,840,  ^d^unweighted n=1,760. | | | |
| ^e^Unweighted n=441, ^f^unweighted n=261, ^g^unweighted n=180. | | | |
| ^h^Unweighted n=128, ^i^unweighted n=56, ^j^unweighted n=72. | | | |

| **Supplemental Table 3** | | | | |
| --- | --- | --- | --- | --- |
| **IFT and the risk of PDS among 2007-2010 PRAMS-MA mothers, by plurality** | | | | |
|  | **% within IFT category^a^** | **% within IFT category who reported PDS^a^** | **unadjusted RR**  **(95% CI)** | **multivariable RR**  **(95% CI)** |
| No IFT (N=3,217) | 88.6 | 9.3 | 1.00 (referent) | 1.00 (referent) |
| Any IFT (N=372) | 11.4 | 11.4 | 1.11 (0.84, 1.46) | 1.22 (0.92, 1.61)^b^ |
| FD | 5.9 | 14.8 | 1.21 (0.92, 1.61) | 1.22 (0.76, 1.98)^c^ |
| DI/IUI | 2.5 | 14.7 | 1.34 (0.81, 2.22) | 1.25 (0.66, 2.35)^c^ |
| ART | 4.8 | 10.0 | 0.98 (0.62, 1.55) | 1.00 (0.61, 1.63)^c^ |
| Mothers with singleton births (N=3,512) |  |  |  |  |
| No IFT | 90.0 | 9.1 | 1.00 (referent) | 1.00 (referent) |
| Any IFT | 10.0 | 12.3 | 1.16 (0.86, 1.54) | 1.26 (0.94, 1.69)^b^ |
| FD | 5.2 | 15.8 | 1.27 (0.87, 1.86) | 1.25 (0.74, 2.10)^c^ |
| DI/IUI | 2.0 | 15.6 | 1.50 (0.87, 2.59) | 1.38 (0.67, 2.82)^c^ |
| ART | 3.8 | 11.7 | 1.00 (0.60, 1.68) | 0.99 (0.57, 1.72)^c^ |
| Mothers with multiple births (N=88) |  |  |  |  |
| No IFT | 37.6 | 22.6 | 1.00 (referent) | 1.00 (referent) |
| Any IFT | 62.4 | 6.4 | 0.40 (0.16, 0.99) | 0.51 (0.21, 1.27)^b^ |
| FD | 31.2 | 9.2 | 0.55 (0.17, 1.77) | 0.91 (0.27, 3.04)^c^ |
| DI/IUI | 21.8 | 11.7 | 0.52 (0.13, 2.09) | 0.70 (0.19, 2.66)^c^ |
| ART | 38.6 | 3.8 | 0.53 (0.19, 1.51) | 0.71 (0.23, 2.30)^c^ |
| ^a^Population-based frequencies were weighted by race/ethnicity. | | | | |
| ^b^Adjusted for maternal age, education, income, mode of delivery, multiple gestations and neonatal risk factors. | | | | |
| ^c^Adjusted for presence of other IFT treatment groups and maternal age, education, income, mode of delivery, multiple gestations and neonatal risk factors. | | | | |
